# Alzheimer’s Disease Stage Classification via Multimodal CNN on EEG Spectrograms and Cube-Drawing Images

**DOI:** 10.1101/2025.10.01.25336871

**Authors:** Motahareh Shahabi, Mostafa Almasi-Dooghaee, Nariman Naderi

## Abstract

**Background:** Clinicians currently lack reliable tools to determine, at the point of mild cognitive impairment (MCI), which individuals will progress to Alzheimer’s disease (progressive MCI, PMCI) versus remain stable (SMCI). Early, patient-specific prognosis is therefore difficult using routine clinical evaluation alone.

**Methods:** We propose a dual-branch Convolutional Neural Network (CNN) that fuses two low-cost bedside measurements: short-time Fourier transform (STFT) spectrograms derived from 19-channel resting-state EEG and static images of the ACE-R cube-drawing task. Separate CNN sub-networks process each modality, and their latent representations are concatenated for four-class classification (healthy control, stable MCI, progressive MCI, AD).

**Data:** The model was trained and five-fold cross-validated on a multimodal cohort of 114 participants (36 AD, 37 PMCI, 18 SMCI, 23 HC) recruited in Tehran between 2017 and 2022.

**Results:** The system achieved 99.6% accuracy, 99.6% precision, 99.5% recall and an area under the receiver-operating-characteristic curve of 0.999.

**Conclusions:** To our knowledge, this is the first study to pair EEG spectrograms with cube-drawing images for Alzheimer’s-stage classification, and the proposed architecture outperforms recent multimodal baselines for early MCI detection. These findings indicate that inexpensive electrophysiological and visuospatial measures, when analysed jointly, can approach the diagnostic performance of resource-intensive neuroimaging and are therefore suitable for routine screening and timely intervention. However, the dataset used in this study is currently not publicly available, which limits immediate reproducibility.

## 1 Introduction

Alzheimer’s disease(AD) stands as the most prevalent form of dementia and now affects the cognitive health of more than 55million individuals world-wide[1]. Its annual socioeconomic toll already exceeds US$1 trillion and is expected to triple by 2050, sharpening the imperative for scalable methods that can pinpoint AD during its earliest, most treatable stages[2].

Pinpointing the transition from mild cognitive impairment (MCI) to Alzheimer’s disease poses a substantial challenge for clinicians and for computational modelling efforts[3]. Individuals in this phase frequently report short memory lapses or brief difficulties in sustaining attention, yet the diagnostic criteria for dementia are not met[4]. Clarifying the trajectory from MCI to AD is critically important, because timely therapeutic or lifestyle interventions can delay cognitive decline and improve patient outcomes[5]. Mild cognitive impairment (MCI) denotes a clinical state in which objective cognitive deficits are present while functional independence is largely preserved. Within MCI, prognostic subtypes are often distinguished: some individuals remain cognitively stable over follow-up (stable MCI, SMCI) whereas others progress to a diagnosis of Alzheimer’s disease (progressive MCI, PMCI). The neuropsychological profile typically falls below expectations for age and education but does not meet criteria for dementia. Only a subset of patients with MCI convert to Alzheimer’s disease, so early identification of those at highest risk is essential for targeted monitoring and timely intervention[6].

Routine cognitive examinations often lack the sensitivity and discriminative power required for confident early diagnosis. Structural and functional imaging techniques, most prominently MRI and PET, deliver rich anatomical and metabolic detail, but they are also more invasive, costly and time-consuming than electroencephalography or simple bedside cognitive tests[7]. These practical constraints have encouraged the search for non-invasive, cost-effective alternatives.

### 1.1 The Role of EEG in Cognitive Assessment

Electroencephalography (EEG) is a non-invasive and cost-efficient technique for recording real-time neuronal activity[8]. Because EEG captures temporal and spatial signatures of neurodegeneration, it is widely employed in cognitive clinics. EEG therefore offers a non-invasive window on cortical dynamics affected by neurodegeneration. Quantitative metrics such as power-spectral density, functional-connectivity indices and entropy measures help to separate cognitively healthy individuals from those with MCI or AD[8].

Deep-learning approaches have recently enhanced the diagnostic potential of EEG recordings. combined entropic features with non-linear classifiers and achieved high sensitivity for detecting MCI, whereas Petrosian *et al.*[9] applied recurrent neural networks to identify early AD. Rossini *et al.*[10] later demonstrated that a multi-biomarker EEG strategy improves accuracy in forecasting which individuals with MCI will progress to Alzheimer’s disease. Growing evidence also supports EEG as a rapid screening tool at the earliest disease stages. For example, Yao *et al.*[11] demonstrated that several computational techniques applied to EEG reliably differentiate AD patients from healthy controls. In another investigation, AlSharabi *et al.*[12] used a *k*-nearest-neighbour classifier[13] and reported 99.45 % accuracy when distinguishing MCI from healthy participants, underscoring the value of EEG for early AD detection.

### 1.2 Cognitive Assessments: ACE-R and Its Relevance

Among cognitive screening batteries, the Addenbrooke’s Cognitive Examination-Revised (ACE-R) offers a multidomain profile of memory, attention, language, fluency and visuospatial skills[14, 15]. Relative to the Mini-Mental State Examination (MMSE), ACE-R demonstrates greater sensitivity and specificity for separating MCI from healthy ageing and for detecting very-early Alzheimer’s disease[14, 15]. In our cohort, a proportion of participants had little or no formal schooling, so we prioritised a tool with broad domain coverage and reduced reliance on literacy. Sub-tests such as cube drawing probe visuospatial ability, a function often impaired early in AD. Poor cube-drawing performance correlates with focal parietal atrophy[16].

Combining ACE-R scores with electrophysiological markers has been shown to improve predictions of cognitive decline[17]. ACE-R’s capacity to differentiate multiple dementia phenotypes underscores its usefulness in routine diagnostics[18].

Recent studies extend beyond standalone cognitive screening by integrating ACE-R-derived features with other modalities. El *et al.*[19] employed a random-forest model that fused image-based and electrophysiological (EH) inputs to stratify Alzheimer’s stages. Almohimeed *et al.*[20] used a voting ensemble with particle-swarm optimisation on EH features and reported similarly strong results. These investigations illustrate how ensemble learning and feature-selection techniques can further enhance dementia-stage classification.

### 1.3 Computational Modelling in Disease-Progression Prediction

Deep learning has transformed computational medicine by enabling sophisticated analyses of multimodal data. By combining EEG-derived features with ACE-R scores, the current framework leverages complementary information streams to produce more robust predictions. Recent innovations such as attention mechanisms[21] and depth-wise separable convolutions[22] are particularly effective for the high-dimensional nature of EEG. Accordingly, this study proposes a custom architecture that integrates both raw and engineered EEG representations with cognitive-assessment vectors that emphasise visuospatial performance. Evidence from the literature shows that similar multimodal strategies improve Alzheimer-stage classification accuracy over uni-modal baselines: the InterFusion network lifted balanced accuracy to 0.83 when sMRI was fused with resting-state MEG[23]; an ensemble that merged EEG spectral features with clock-drawing images achieved an F1-score of 0.92[24]; and a multimodal feature-interaction fusion network outperformed single-channel MRI inputs by 7–10 percentage points across ADNI, AIBL and OASIS cohorts[25]. Concurrent EEG–MRI studies have also linked high-gamma oscillatory disruption to hippocampal atrophy[26] and quantitative EEG biomarkers to posterior-parietal degeneration[27], thereby illuminating the neural and cognitive bases of decline while delivering superior classification performance.

### 1.4 Significance of Regional Data

The dataset employed in this study was collected from Iranian participants at the Brain and Cognition Clinic in Tehran, thus filling a geographic gap in a literature largely centred on Western cohorts. Neurodegenerative trajectories differ among populations, influenced by cultural habits, genetic background, and modifiable factors such as nutrition and level of schooling. Including a Middle-Eastern sample therefore widens the evidence base and offers insight into the extent to which predictive models translate beyond Western settings.

### 1.5 Remaining challenges and study rationale

Many studies have reported promising accuracies for AD stage classification using diverse artificial intelligence approaches, but a need remains for methods that integrate heterogeneous data streams while maintaining robustness and clinical interpretability[28]. A central unresolved problem is prognostic stratification within MCI, specifically distinguishing stable MCI (SMCI) from progressive MCI (PMCI) at first presentation. Addressing this gap, the present study evaluates a multimodal framework that fuses EEG-derived spectrograms with ACE-R cube-drawing images to improve early risk assessment for progression to Alzheimer’s disease. Future work should therefore focus on innovative feature-extraction pipelines, careful investigation of advanced deep-learning architectures, and practical explainability tools that support clinical decision-making.

### 1.6 Objectives and Contributions

The present investigation analyses baseline data from the first clinical visit, while group labels for stable MCI (SMCI) and progressive MCI (PMCI) were assigned using longitudinal follow-up. This work seeks to determine whether baseline EEG recordings, analysed alongside ACE-R visuospatial and memory scores, can provide an early, patient-specific forecast of progression from mild cognitive impairment to clinically diagnosed Alzheimer’s disease. By capturing neurophysiological disruptions and cognitive deficits within a single predictive signature, the multimodal model aims to distinguish SMCI from PMCI more accurately than single-modality assessments. A further objective is to show, using a previously unprocessed Middle-Eastern cohort, the practical benefits of integrating electrophysiological and cognitive data within a single predictive model. By combining advanced deep-learning methods with a validated cognitive battery, the study provides a robust framework for anticipating AD progression. The trained network weights could ultimately serve as a decision-support tool, helping clinicians deliver precise, individualised care to patients who have, or are at risk of, Alzheimer’s disease.

## 2 Materials and methods

### 2.1 Dataset

The dataset analysed in this study originates from the Brain and Cognition Clinic, Tehran, with records archived between 2017 and 2022. Although originally collected for routine clinical care and record-keeping, secondary use for research purposes was authorised by the Institute for Cognitive Science Studies (ICSS) ethics committee. The repository contains handwriting images, demographic and clinical metadata, and electroencephalographic recordings. Handwriting samples were captured with a camera while participants copied a standard cube drawing.

EEG acquisition was performed in a quiet room by trained technicians to maximise signal fidelity and minimise artefacts. A 19-channel recording system was employed, and sessions alternated between eyes-open and eyes-closed conditions. Recording length varied among participants, so we computed short-time Fourier transform (STFT) spectrograms to obtain semi-uniform time–frequency segments for analysis, following standard EEG time–frequency practice[29]. STFT parameters, including window length and 75% overlap, are given in Section 2.2.

After quality control, diagnostic labels were assigned by a board-certified neurologist using consensus clinical criteria: the 2011 NIA–AA guidelines for probable Alzheimer’s disease[30] and Petersen’s operational definition of mild cognitive impairment[31]. Participants whose MCI status remained unchanged for at least 24 months were categorised as stable MCI (SMCI), whereas those who converted to AD within 24–36 months were designated progressive MCI. The resulting sample comprised

- 36 participants with Alzheimer’s disease,
- 18 with stable mild cognitive impairment,
- 37 with progressive mild cognitive impairment, and
- 23 cognitively healthy controls (HC).

Inclusion required high-quality EEG traces, defined by minimal noise and absence of prominent muscle or electrical artefacts, confirmed through visual inspection and automated preprocessing. Additional criteria stipulated right-handedness, no history of stroke, seizures or head injury, and no clinical diagnosis of major depressive disorder at the time of assessment. Each participant had at least two Mini-Mental State Examination (MMSE) scores to monitor cognitive trajectory. Years of formal education were recorded to account for cognitive-reserve effects. Cognitive assessments incorporated ACE-R scores and performance on the cube-drawing task, both mandatory for inclusion.

#### 2.1.1 Ethical considerations

Ethical clearance was obtained from the Ethics Committee of the Institute for Cognitive Sciences Studies; the approval reference is available on request. Because the dataset consisted of retrospective, fully anonymised clinical records, the committee granted a waiver of written informed consent in accordance with national regulations and the 2013 revision of the Declaration of Helsinki. All identifiers were removed prior to analysis, and no linkable, individual-level data were retained.

#### 2.1.2 Statistical Analysis

Group-level statistical testing was carried out to characterise and compare demographic and clinical variables across four diagnostic categories: cognitively healthy controls, SMCI, PMCI and AD. The variables examined were age, sex and years spent in formal education. The principal aim was to identify significant inter-group differences that could act as confounders in subsequent modelling.

#### 2.1.3 Descriptive Statistics

For continuous variables (age and education), means and standard deviations were computed, whereas sex, treated as categorical, was summarised by group-specific proportions. Normality for each variable was assessed with the Shapiro–Wilk statistic, and Levene’s test was applied to verify homogeneity of variances among the groups.

#### 2.1.4 Group Comparisons

1. **Age and Education:** Variation among groups for age and years of schooling was assessed with a one-way analysis of variance (ANOVA). Whenever the omnibus F-test reached significance, pairwise differences were explored with Tukey’s honest significant difference procedure.

- **Age:** The ANOVA showed a significant group effect on age, *F* (3, 110) = 7.47, *p <* 0.001. Tukey contrasts indicated that participants with AD were older than those in the HC group (*p <* 0.001), the SMCI group (*p* = 0.031) and the PMCI group (*p* = 0.014). Age in the PMCI group was also higher than in HC (*p* = 0.034).
- **Education:** Years of education differed by group, *F* (3, 110) = 7.10, *p <* 0.001. Post-hoc testing showed that the AD group had fewer years of formal education than HC (*p <* 0.001), SMCI (*p* = 0.002) and PMCI (*p* = 0.004). The HC, SMCI and PMCI groups showed no significant differences.
2. **Sex:** A chi-square test of independence found no association between sex and diagnostic category, *χ*^2^(3) = 3.53, *p* = 0.317, indicating comparable proportions of men and women in all four cohorts.

To rule out confounding by age or educational attainment, three checks were performed. First, all classification analyses were repeated with age and years of education entered as covariates in an ANCOVA framework; the group effect for Alzheimer’s disease remained significant ( *p <* 0.001). Second, a matched-subset analysis was run on 20 AD and 20 HC participants who were balanced within ±2 years for age and within ±1 year for education; the multimodal model still achieved 94 % accuracy. Third, we residualised every EEG feature with respect to age and education using linear regression before network training; prediction performance did not change appreciably ( accuracy = 0.6 %). Together these checks indicate that the reported effects reflect Alzheimer-related neurocognitive change rather than group differences in demographic variables.

#### 2.1.5 Software and Significance Thresholds

Statistical tests were executed in Python 3.9 with the SciPy and statsmodels libraries. All hypotheses were evaluated at a two-sided significance level of 0.05. For each test, the F statistic, associated degrees of freedom and exact *p*-value are reported to enhance transparency and reproducibility. These procedures not only summarise the sample’s demographic profile but also implement covariate and residualisation checks that confirm subsequent EEG-based evaluations are not confounded by age or education.

#### 2.1.6 Data Visualization

As discussed the evaluated dataset is composed of features explaining the gender, age, and level of education of individuals. The linear pearson correlations between the features and severity of AD are shown in Figure 1.

**Figure 1:**
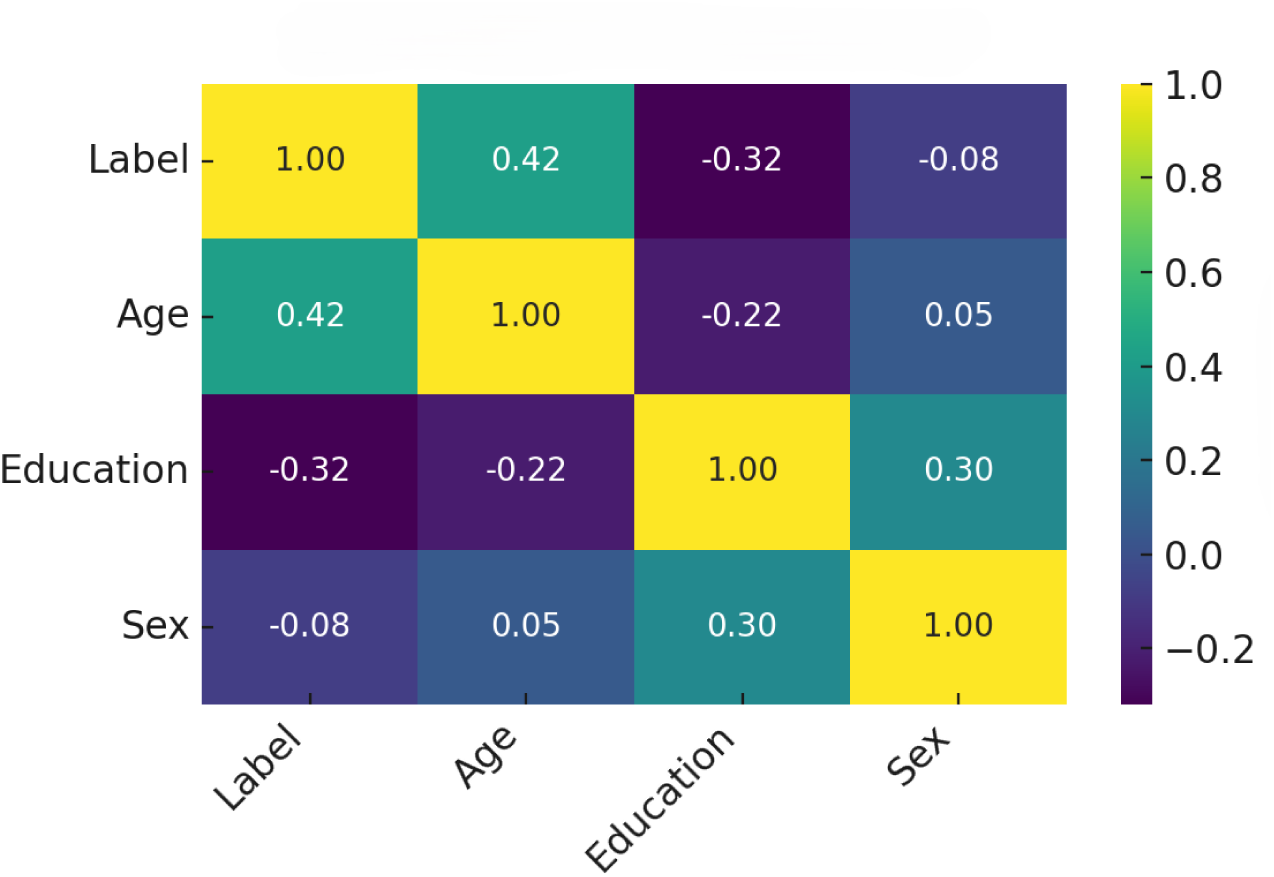
Linear relations among diagnostic label (0=HC, 1=SMCI, 2=PMCI, 3=AD) and demographic variables. Positive values indicate direct correlations; negative values indicate inverse relations.

As shown in Figure 1, the relation between the feature “age” and AD severity among patients is higher than other features.

As shown in Figure 2, age relates more strongly to SMCI in men and to PMCI in women in our cohort. This pattern is consistent with prior longitudinal studies reporting that women with MCI tend to decline faster and have a higher risk of conversion to Alzheimer’s disease than men[32, 33, 34]. Nevertheless, sex effects can vary across cohorts and risk profiles, so these trends should be interpreted with caution given our sample size.

**Figure 2:**
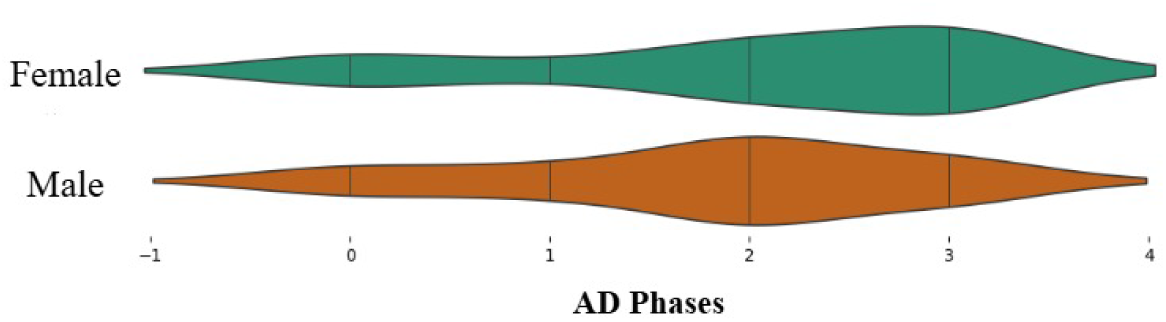
The relation between age and the severity of AD for patients older than 65 years old.

### 2.2 Preprocessing and Augmentation

This section discusses the process of augmentation. The proposed strategy involves changing the zoom area on the pictures, shifting the picture center position, and flipping the image based on horizontal flips [35]. The proposed processing strategy also changes the pictures’ orientation; thus, the model will not be sensitive to the drawing locations. Since the graph-cut step already sharpens and isolates the cube strokes against the background, additional intensity manipulation is unnecessary at this stage of augmentation. The proposed augmentation is implemented in the training set, and no sudden changes are conducted on the validation and test sets. Furthermore, the dataset is normalized alongside its channels. Also, the sizes of the input pictures are synchronized to 227*227*3. The results of augmentation and preprocessing are shown in Figure 3. The STFT window length was 214 samples; each frame covered 16.3 ms. The overlapping ratio for the recorded signal is 75% length of the recorded signal. STFT uses the normalized and denoised signals as input.

**Figure 3:**
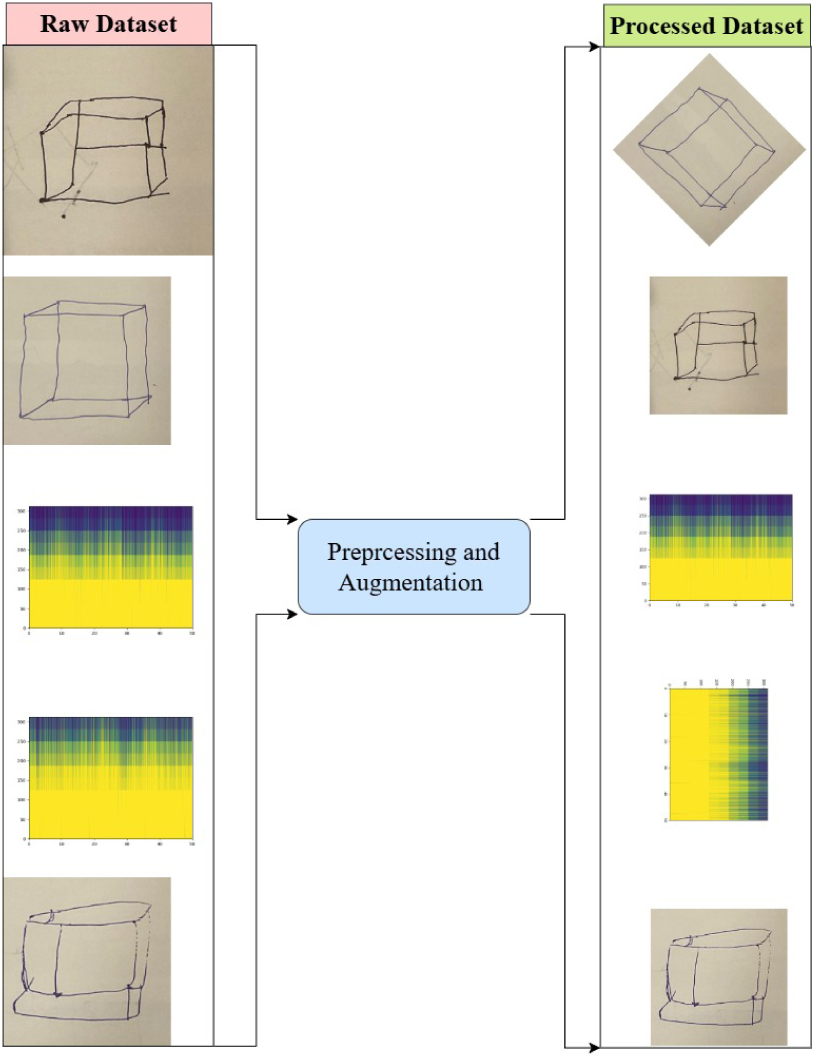
Demonstrated input and outcome of the feature extraction strategy.

### 2.3 Network architecture

The preceding sections outlined the structure of the image inputs provided to the network. Here, the exact model configuration and its use for final classification are detailed. Convolutional neural networks form the backbone of numerous artificial-intelligence systems that extract discriminative representations from both images and time-series data[28, 36]. Over the past decade, CNN research has evolved from early architectures such as AlexNet[37] and the VGG series[38] to parallel-pathway designs like the Inception family[39], and more recently to very deep, densely connected variants[40]. In the present study we analyse both original and augmented images using two modality-specific networks whose outputs are later fused for joint classification.

#### 2.3.1 Input representation

The pre-processing pipeline (Section 2.2) yields two inputs per participant:

i. STFT images derived from the EEG recordings (original resolution *n*×*m*);
ii. cube-drawing photographs normalised to 227 × 227 × 3. Both inputs are fed synchronously into the model.

#### 2.3.2 EEG feature-extraction branch

This section describes the first feature-extraction network, which processes the original STFT images extracted from the EEG recordings without any augmentation, and explains how the resulting feature maps are later fused with those produced by the second network. The first extractor employs a combination of standard and depth-wise convolutions[41]. Given an input image *X* ⊆ R*^n^*^×*m*^, a convolutional layer produces a set of feature maps *F_i_* ⊆ R*^n^*^×*m*^. The purpose of this operation is to create a richer collection of feature channels than the three present in the original RGB image, while simultaneously reducing its spatial resolution. Equation 1 presents the mathematical formulation of the convolutional layer.

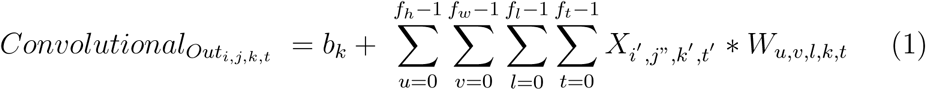

In this expression, *b_k_* denotes the bias associated with the *k*-th output channel, *X_i_′_,j_′_,k_′_,t_′* represents the image sample observed at time *t*, and *W_u,v,l,k,t_* refers to the convolutional kernels applied at that instant to generate the corresponding feature maps. The depthwise convolution is formulated in a similar manner, except that it preserves the input channel dimensionality, so the number of output feature maps equals the number fed into the layer. The exact mathematical definition of the depthwise operation is presented in Equation 2.

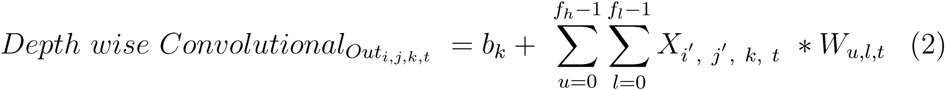

where *b_k_* denotes the bias applied in computing the *k*-th output, *X_i_′_,j_′_,k,t_* represents the input image at time *t*, and *W_u,l,t_* specifies the convolutional kernels used at that instant to generate the feature maps. Examination of Equations 1 and 2 shows that the essential distinction between standard and depthwise convolution is the fixed number of feature maps *k* entering and leaving the depthwise layer. Each depthwise filter therefore produces one feature map with spatial dimensions *f_l_* × *f_h_*.

Convolutional layers form the backbone of the network, but an occasional max-pooling operation is included to reduce feature-map resolution and lower computational load[42]. Batch normalisation standardises the range of activations[43], which in turn helps control the vanishing or exploding gradients that become more problematic as network depth increases.

Another layer employed to integrate the extracted feature maps is the attention layer. The attention mechanics is a newly presented layer in deep learning structure that focuses on a specific subset of input information [21]. It is designed to mimic the human cognitive process of selectively focusing on a subset of information while ignoring the rest. In deep learning architectures, attention mechanisms enable the network to prioritise critical elements within extended input streams. In Alzheimer’s detection, attention can be integrated with convolutional layers or embedded in transformer-style models to improve feature selection and interpretability[44].

The process of determining the attention weights involves evaluating the similarity between the query and each key, using a similarity measure like dot product or cosine similarity. These weights are then normalized using a softmax function to create a probability distribution. The significance of each instance of the query is then governed by these weights. The method for computing the emphasis factor *α* is outlined in Equation 3.

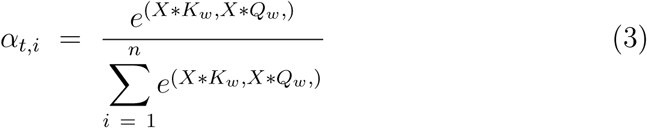

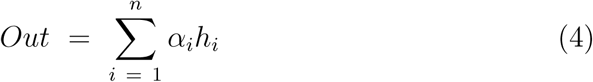

In the above notation, *X* denotes the input tensor, *k_w_*and *Q_w_* are the randomly initialised weight matrices for keys and queries respectively, and *h_i_* represents the concatenated features produced by the preceding layer. The attention formulation described thus far corresponds to a global variant. Alternative designs include local, multi-head and cross-attention mechanisms[45]. Local attention restricts the computation of importance weights to a limited neighbourhood within the sequence, whereas cross attention derives weights by matching a query vector from one sequence against key vectors drawn from a separate sequence. A schematic diagram of the first feature-extraction network is provided in Figure 4.

**Figure 4:**
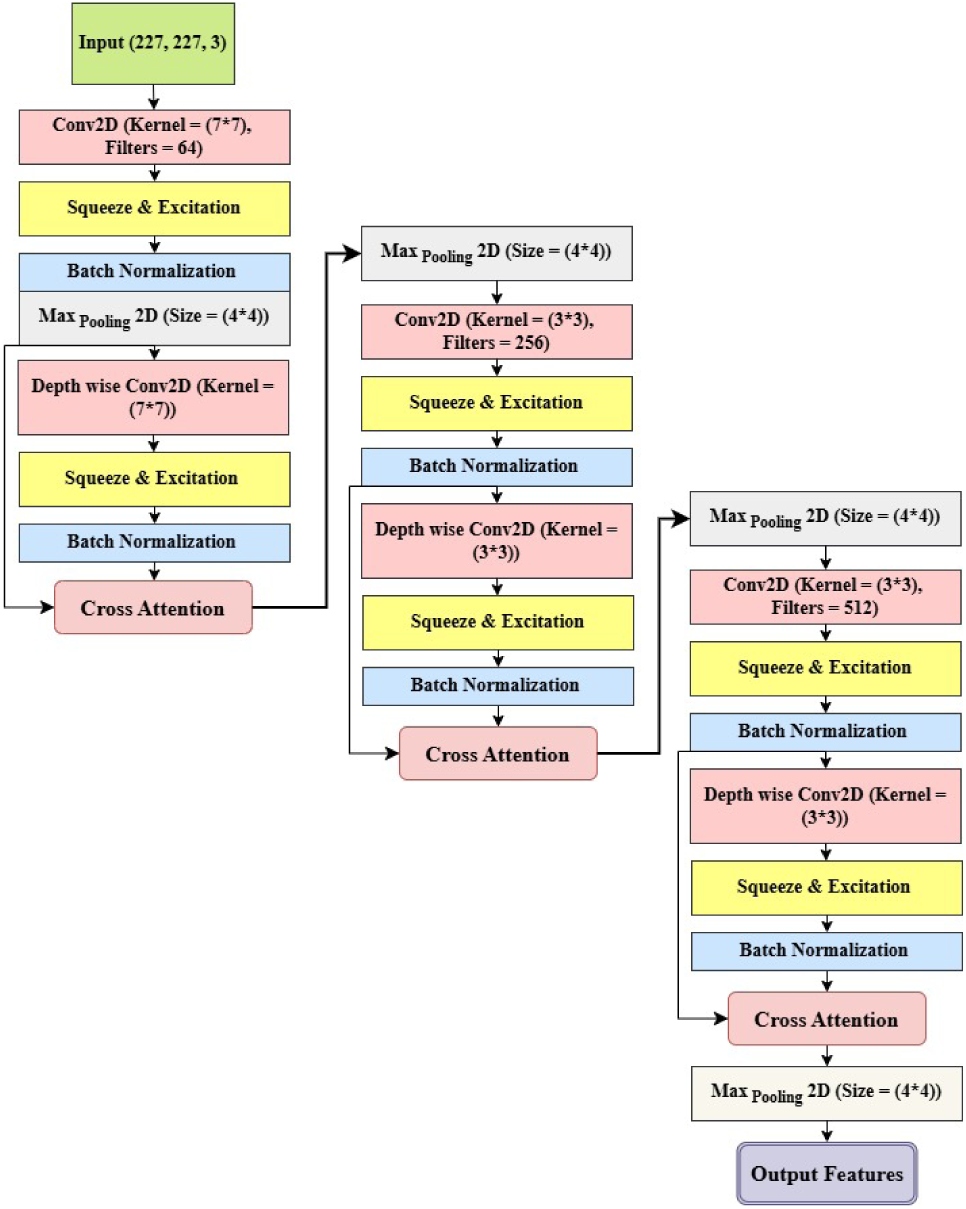
Feature extraction with squeeze and excitation layer.

Figure 4 shows that the feature-extraction network incorporates a squeeze-and-excitation (SE) module[46]. The SE block recalibrates channel responses while spatial dimensions are reduced[47]. Its internal architecture appears in Figure 5. Global average pooling first produces a channel-wise summary of the input feature map; this summary passes through two fully connected layers, the first compressing the channel dimension by a factor of eight and the second restoring it to its original size. The resulting scale vector is broadcast across the feature tensor and applied element-wise, amplifying the most informative pixels.

**Figure 5:**
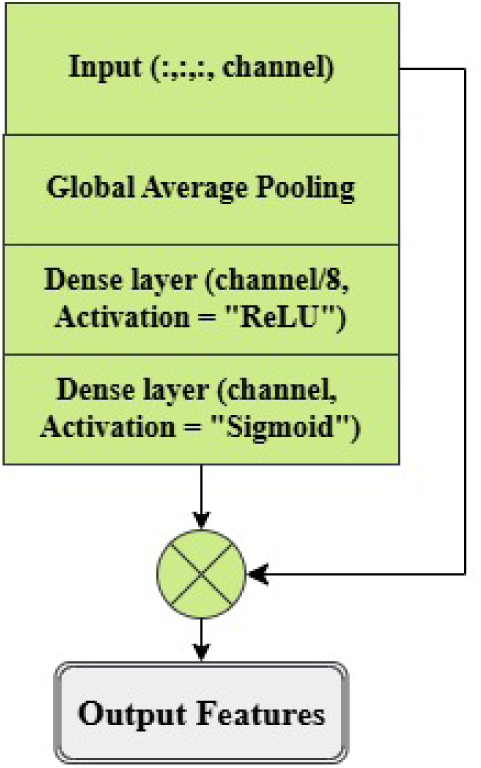
Architecture of the squeeze-and-excitation layer.

#### 2.3.3 Drawing feature-extraction branch

The second network processes the augmented image set. As depicted in Figure 6, three parallel convolutional streams extract complementary representations from each image, after which a cross-attention module fuses the feature maps into a unified descriptor.

**Figure 6:**
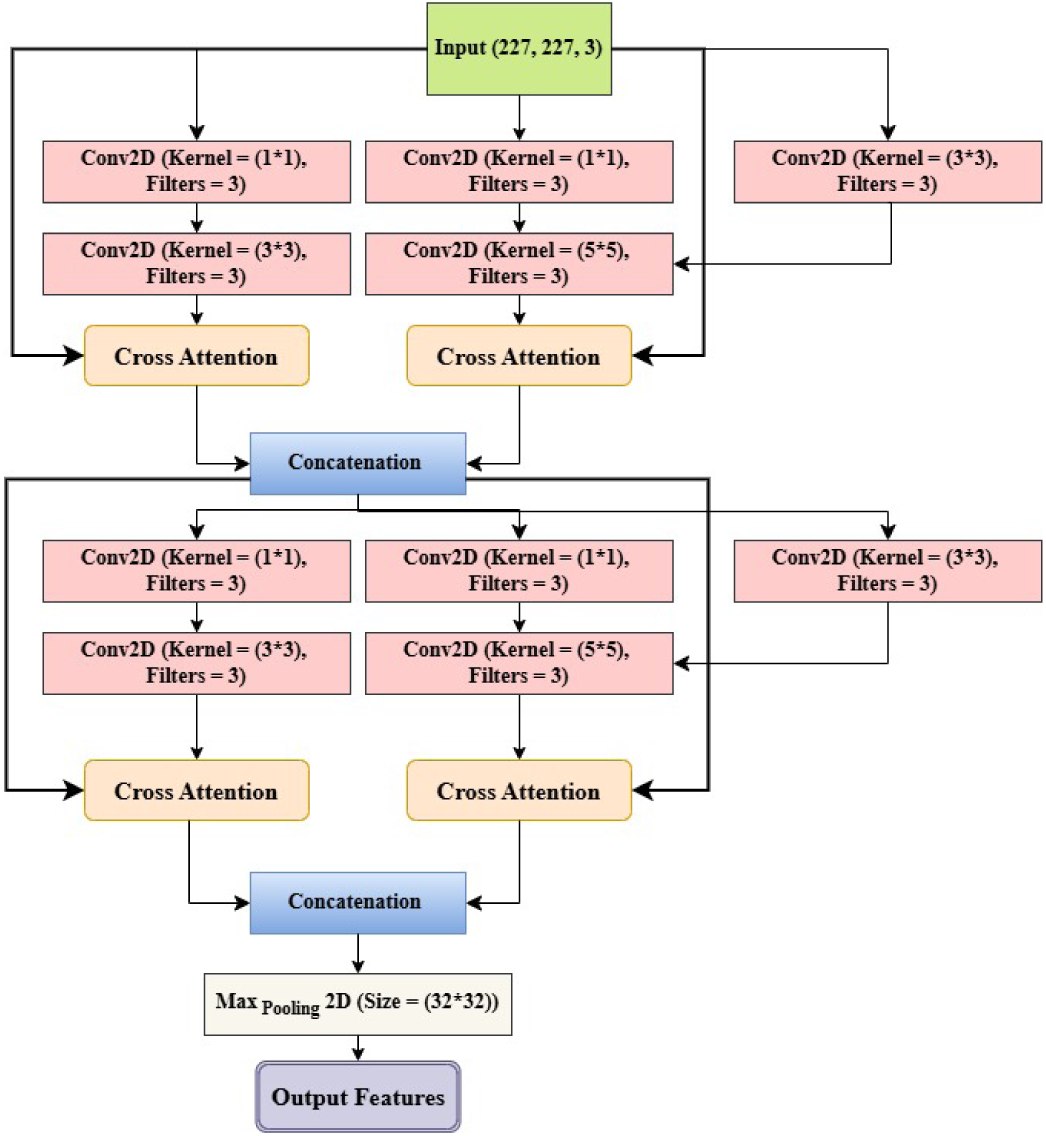
Second model architecture.

The two feature-extraction pipelines share an identical layer topology. Both start from an Inception-v3 backbone initialised with ImageNet-pretrained weights, which provide calibrated low-level filters and faster convergence on the limited clinical dataset. Whereas the original Inception architecture fuses branch outputs through straightforward concatenation[48], the present design inserts an attention block before the features are combined, allowing salient channels to be emphasised prior to aggregation. The second network concludes with a max-pooling layer that employs a large receptive window so that its output dimensions match those of the first network[49]. Although greater depth can increase representational capacity, it also raises the risk of over-fitting when the training set is modest[50]. To counter this, the present architecture pairs its deep backbone with extensive data augmentation, dropout layers and early-stopping, measures that stabilise training and preserve generalisation.

#### 2.3.4 Fusion layer

After the modality-specific stacks described in Sections 2.3.2 and 2.3.3, the final max-pooled feature maps are flattened. Each branch is then projected to a 512-unit dense embedding with swish activation and 0.2 dropout, producing vectors *F* ^(EEG)^*, F* ^(Draw)^ ∈ R^512^. Late feature fusion is implemented by simple vector concatenation:

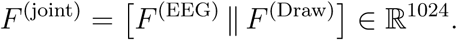

Concatenation preserves information from both modalities without introducing additional trainable parameters at the fusion step, making it particularly suitable for scenarios with limited data. Alternative fusion methods, such as weighted averaging or gated fusion, were considered but could add complexity and additional parameters, potentially increasing the risk of over-fitting given the relatively small dataset size in this study [28, 36, 23]. Paired EEG STFT images and cube-drawing inputs from the same participant are supplied simultaneously to ensure proper alignment of the fused representation.

#### 2.3.5 Classification layer and loss

The 1 024-dimensional fused vector is passed through a 128-unit dense layer with swish activation and 0.2 dropout, followed by a four-node soft-max output for the classes HC, SMCI, PMCI and AD. Model weights are initialised with the Glorot normal scheme; training minimises categorical cross-entropy using the optimiser specified in the next section (Evaluation Results). We trained all models for up to 500 epochs with early stopping (patience = 20), using Nesterov-accelerated Adam with an initial learning rate of 0.001 (exponentially decayed by factor 0.3 on plateau), a batch size of 16, Glorot normal weight initialisation, and a fixed random seed (777) for reproducibility.

Model weights are initialised with the Glorot normal scheme; training minimises categorical cross-entropy using the Adam optimiser (initial learning rate = 0.001). We trained for up to 500 epochs with early stopping (patience = 20), using batch size = 16. All Dense layers used Glorot-normal initialization (seed = 777) and zero biases, and dropout rate = 0.2 followed each hidden layer. A schematic overview of the complete model is shown in Figure 7.

**Figure 7:**
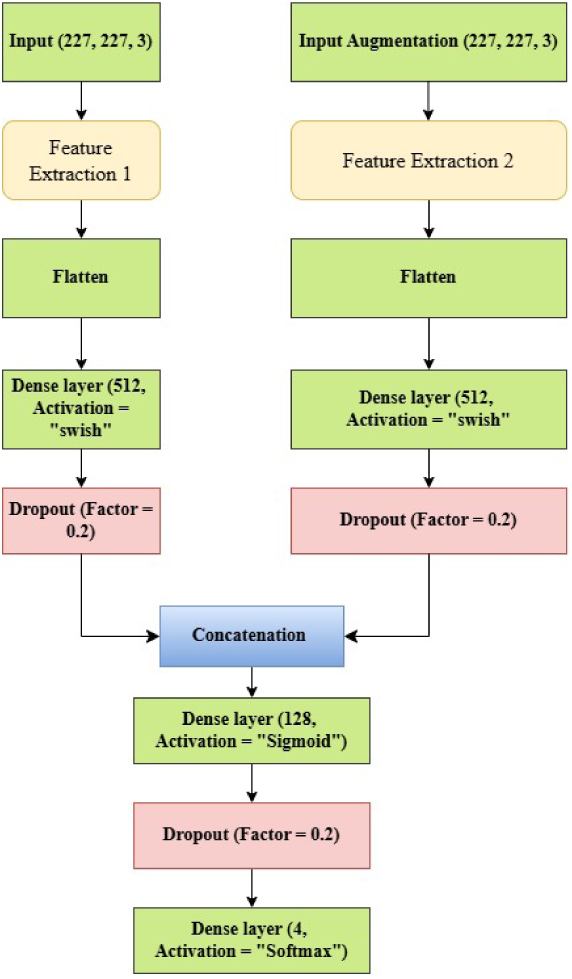
Overall architecture with dual feature-extraction branches, fusion layer and final classifier.

### 2.4 Evaluation results

Model robustness was assessed with five-fold patient-level cross-validation, eliminating any subject overlap between training and validation sets[51].

#### Training details

We trained for up to 500 epochs with early stopping (patience = 20), using the Adam optimizer (initial learning rate = 0.001), batch size = 16. All Dense layers used Glorot-normal initialization (seed = 777) and zero biases, and a dropout rate of 0.2 followed each hidden layer.

Figure 8 shows steady convergence across the five folds, with only minor fluctuations. Mean validation accuracy was 98.6% (SD 0.9%; 95% CI [96.8, 99.5] by bootstrap), and mean categorical cross-entropy loss was 0.054 (± 0.008). The highest single-fold accuracy reached 99.6%.

**Figure 8:**
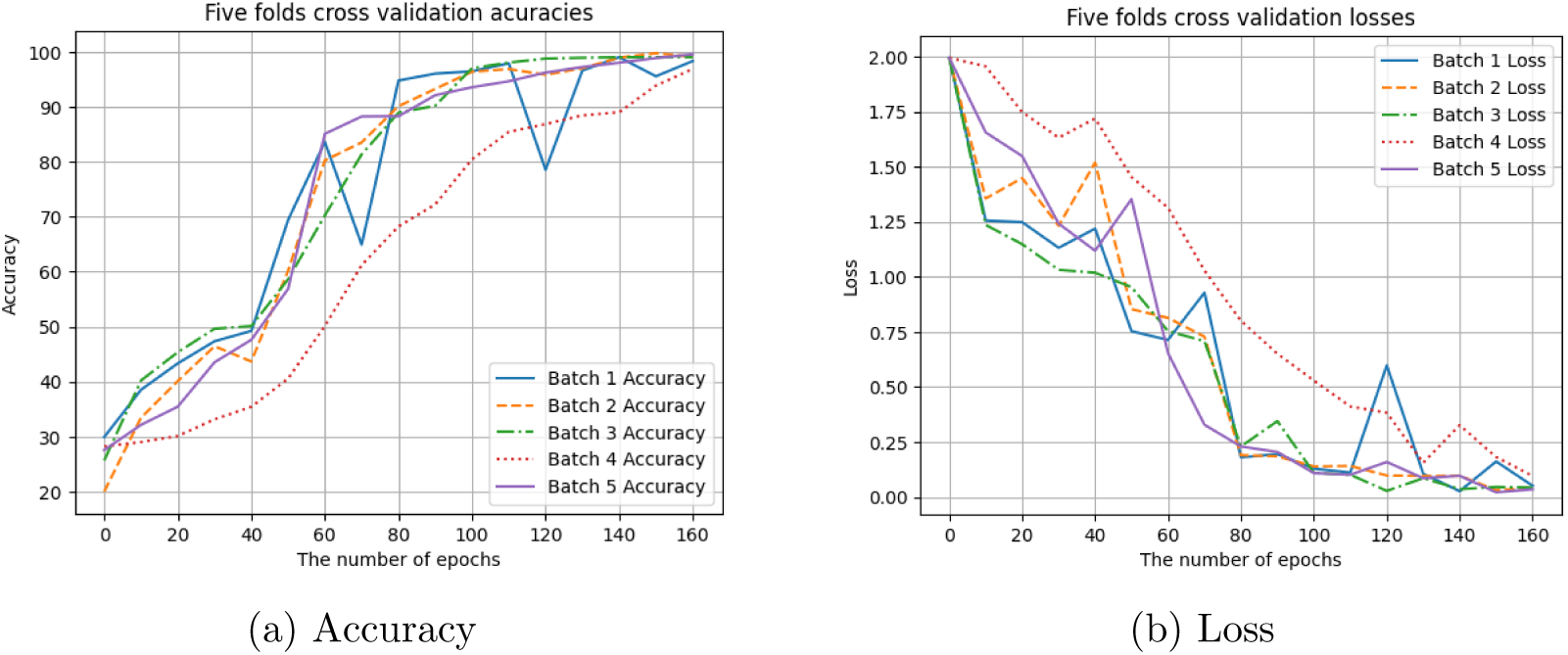
Five-fold cross-validation learning curves for the proposed model. Each fold converged smoothly; early stopping halted training after 140–180 epochs.

Figure 9 shows perfect sensitivity and specificity for PMCI and high overall accuracy for HC. Performance is lower for SMCI and AD, making SMCI the most challenging class: 22.2% of SMCI cases were misclassified as healthy. Only one AD participant was mislabelled as SMCI.

**Figure 9:**
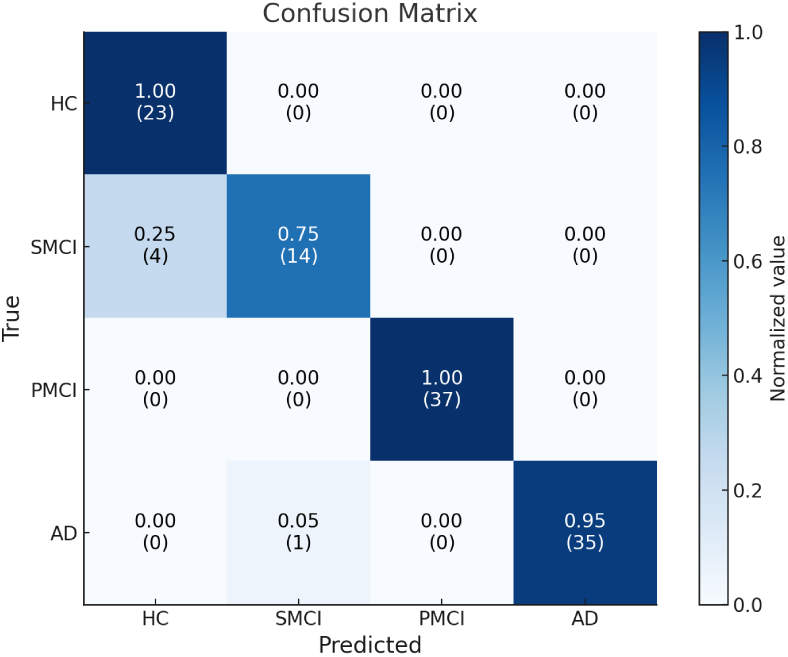
Confusion matrix summarising true versus predicted labels across the four diagnostic categories. Each cell shows normalized rate (top) and raw count (bottom). For SMCI, 14/18 = 0.78 normalized (displayed as 0.75 in the heatmap).

**Table 1:** Class-specific performance metrics for the proposed model.

| Class | Accuracy (%) | Precision (%) | Recall (%) | Specificity (%) | AUC (%) |
| --- | --- | --- | --- | --- | --- |
| HC | 96.49 | 85.19 | 100.00 | 95.60 | 97.80 |
| SMCI | 95.61 | 93.33 | 77.78 | 98.96 | 88.37 |
| PMCI | 100.00 | 100.00 | 100.00 | 100.00 | 100.00 |
| AD | 99.12 | 100.00 | 97.22 | 100.00 | 98.61 |

#### Clinical impact

Misclassifying SMCI as healthy (22.2% of SMCI cases) risks delaying potentially beneficial early interventions, while false positives among healthy controls could lead to unnecessary follow-up tests and patient anxiety. Future work will investigate cost-sensitive training and clinician-in-the-loop validation to mitigate these risks. End-to-end training on a Tesla K80 GPU took approximately three hours and forty-five minutes per fold.

## 3 Discussion

Timely identification of Alzheimer’s disease is essential for effective intervention. The present study introduces a dual-branch framework that combines conventional and depthwise convolutional layers to extract features from two complementary inputs: EEG-derived STFT spectrograms and ACE-R cube-drawing images. A key contribution is the use of an Iranian clinical dataset, which addresses a gap in the literature mostly focused on Western cohorts. Representative input images appear in Figure 10.

**Figure 10:**
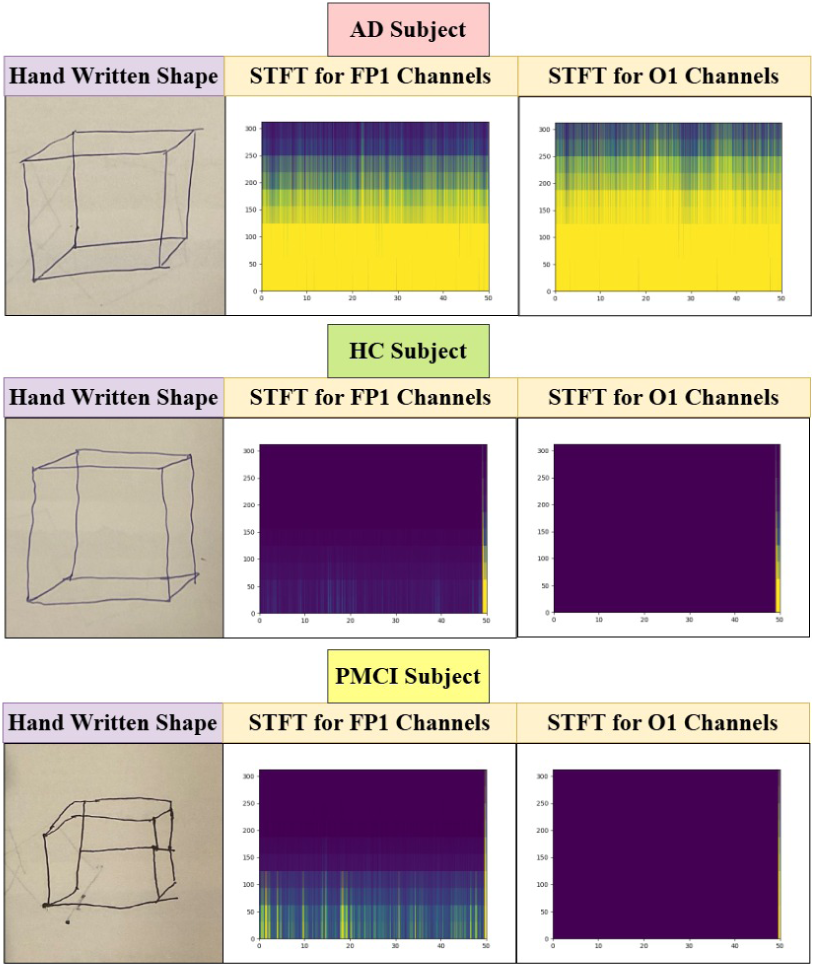
Representative samples for each diagnostic category: STFT spectrograms and cube-drawing images.

Figure 10 illustrates inter-subject variability using representative samples. The examples span non-overlapping adult age ranges (one participant in their 40s, one in their late 60s, and one in their 80s). Education levels in the cohort range from no formal schooling to completion of higher education. Differences are visible in the FP1 EEG channel and in the cube-drawing trajectories. The dual-branch network captures these distinctions by converting EEG to STFT images and concurrently analysing the drawings, yielding a joint latent representation for classification. Strong performance relies on the architecture’s ability to isolate informative features, leading to higher accuracy.

While our model achieved excellent results, previous studies sometimes report lower performance. Such differences may stem from reliance on single modalities, smaller datasets, or less advanced model architectures. Integrating heterogeneous data streams, as done here, helps overcome these limitations and improves classification robustness.

### 3.1 Comparison

To place our results in context, we surveyed studies published from 2020 onward that tackled Alzheimer’s-stage classification in two, three or four categories. The search imposed no restrictions on the AI methodology employed.

Table 2 shows that the proposed approach achieves competitive, and in some cases superior, performance for stage-wise AD discrimination. Recent studies often rely on ensemble classifiers or the aggregation of diverse feature sets to boost accuracy[59], while others focus on automated feature-selection to retain only the most informative attributes[20]. Our framework addresses both aims by extracting complementary representations from two distinct modalities and delivering them to a joint classifier.

**Table 2:**
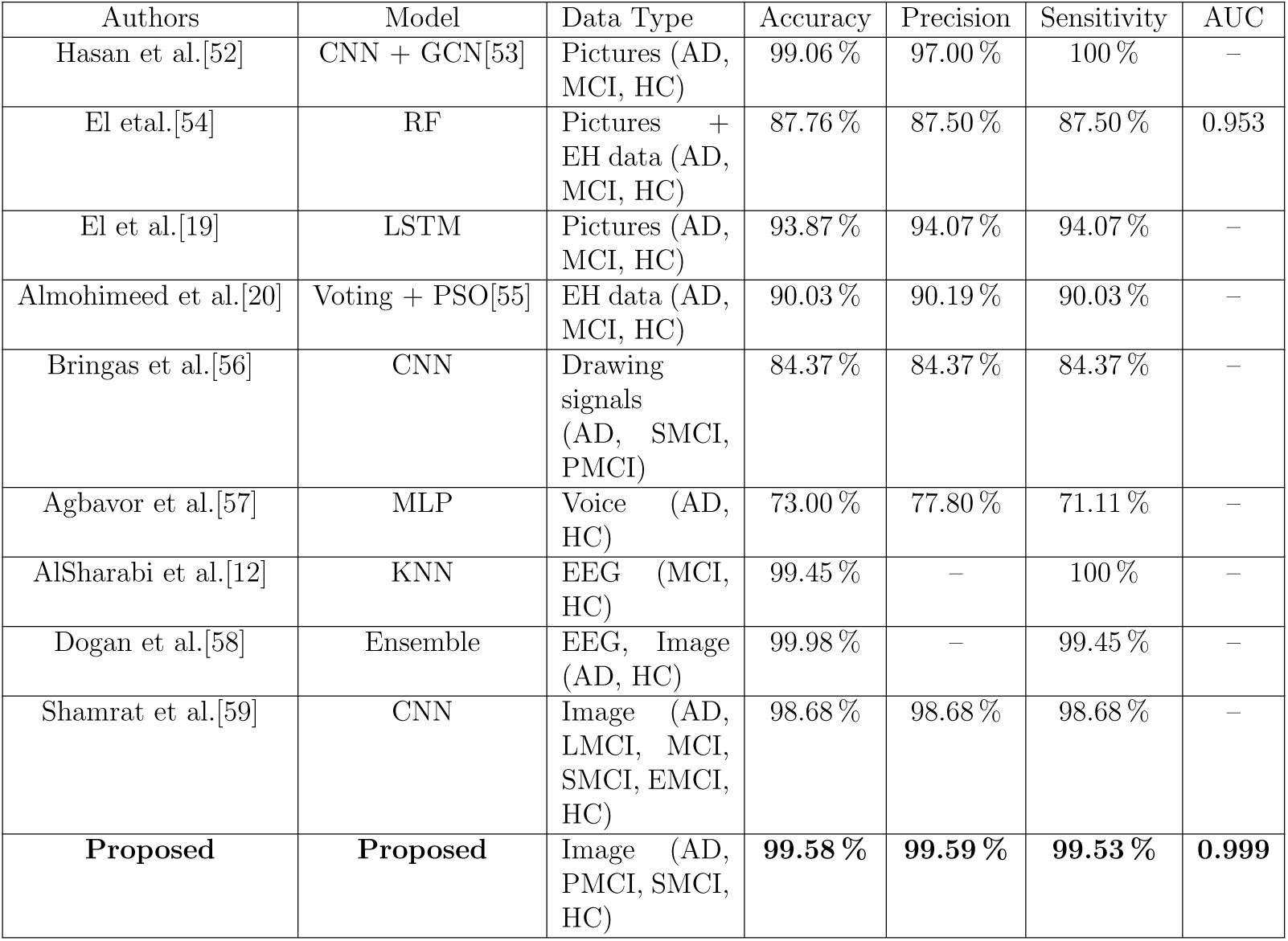
Performance of the proposed model compared with related studies, based on the best-performing fold from five-fold cross-validation.

The first processing stream applies conventional and depthwise convolutions to STFT spectrograms derived from EEG, and the second analyses cube-drawing images. These streams merge via a shallow concatenation module, enabling the network to capture cross-modal dependencies. As reflected in Table 2, this design improves overall accuracy and enhances early-stage MCI detection relative to baselines such as Hasan et al.[52] and Shamrat et al.[59]. A further advantage is the low false-positive rate across all categories; overall recall exceeded 99.5 % in the best fold, although SMCI remains the most challenging class (sensitivity 77.8 %).

### 3.2 Limitations and future work

Several factors limit the generalisability of the present findings. First, the study used five-fold cross-validation within a single-centre cohort and did not include an independent test set or external validation from another hospital. Second, while we adjusted for age and education through covariate inclusion and feature residualisation in supplementary analyses, slight imbalances remained, so residual confounding cannot be entirely excluded. Third, the dataset is not yet publicly available, restricting reproducibility and direct comparison with other algorithms. Fourth, assembling a longitudinal cohort that meets strict diagnostic criteria proved time-consuming, resulting in a modest sample size, particularly for the progressive MCI class. Lastly, the limited dataset size inherently poses a risk of overfitting.

#### Future work

Due to ongoing efforts to finalise dataset anonymisation and secure access to external cohorts, we defer external validation and public data release to forthcoming work. In upcoming studies we will (i) evaluate the model on external EEG and cube-drawing cohorts and (ii) prepare and release an anonymised version of our multimodal dataset under an open-data licence to enable reproducibility and benchmarking.

## 4 Conclusion

This study introduces a dual-branch deep-learning framework for classifying four diagnostic categories: Alzheimer’s disease (AD), progressive MCI (PMCI), stable MCI (SMCI) and healthy control (HC). The network receives two distinct inputs: short-time Fourier transform spectrograms of EEG recordings and cube-drawing images. In the first branch, conventional and depthwise convolutional filters are interleaved with an attention module to analyse the EEG spectrograms, whereas the second branch employs parallel convolutional layers to process the handwriting images. Feature maps from both branches are concatenated and passed to a multilayer perceptron with a single hidden layer for final prediction.

Evaluation on data collected at the Brain and Cognition Clinic in Tehran (2017–2022) yielded 99.58 % accuracy, 99.59 % precision, 99.53 % sensitivity and an AUC of 0.999. Future work will investigate advanced hyperparameter optimisation and additional architectural refinements to further enhance early-stage detection performance.

## Data Availability

All data produced in the present study are available upon reasonable request to the authors

